# Covid-19 Epidemic Prediction in France : the Multimodal Case

**DOI:** 10.1101/2021.10.09.21264794

**Authors:** J.-P. Quadrat

## Abstract

In two previous papers we have proposed models to estimate the Covid-19 epidemic when the number of daily positive cases has a bell shaped form that we call a mode. We have observed that each Covid variant produces this type of epidemic shape at a different moment, resulting in a multimodal epidemic shape. We will show in this document that each mode can still be estimated with models described in the two previous papers provides we replace the cumulated number of positive cases *y* by the cumulated number of positive cases reduced by a parameter *P* to be estimated. Therefore denoting *z* the logarithm of *y −P, z* follows approximately the differential equation ż = *b −az*^*r*^ where *a, b, r* have also to be estimated from the observed data. We will show the obtained predictions on the four French modes April, November 2020, May and September 2021. The comparison between the prediction obtained before the containment decisions made by the French government and the observed data afterwards suggests the inefficiency of the epidemic lockdowns.

## 1 Introduction

The forecast of the spread of the Covid epidemic is very important to take governmental decisions such as containment policies. In [2] we have shown that time-invariant SIR models [5, 4] were not effective to forecast the number of contaminated-detected people and we have proposed a new model. It was a dynamical linear model forecasting the logarithm of the cumulated infected-detected population *z*. In [2] The dynamics was *ż* = *b − az*. In [3] we have shown that this model could be improved using the dynamics *ż* = *b −az*^*r*^ where *r* is a real number to be estimated from the observed data. These models are good for what we call an unimodal epidemic, which is a daily number of contaminated people with a bell shaped form. But it is clear now that the Covid epidemic is a succession of different epidemics coming from different variants and therefore the epidemic may be multimodal. We propose here a method to estimate the different modes. Remarking that each mode corresponds to the domination of one variant it is natural to think that the previous models applied not to *y* but to *y − P* is valid during the time period of domination of the considered variant. Then *P* has the interpretation of the cumulated positive cases number due to previous variants. The model being effective only during the period of domination of the considered variant. The difficulty is to estimate this parameter *P* We will explain a way to estimate the four parameters *a, b, r, P* and will give the results obtained for the four French modes.

Based on these models able to predict the different modes, we can judge the effect of the three French confinements by comparing the predictions with the observed data. It appears that the model can predict correctly the mode based on the data known before the confinement decision. This fact suggests that the confinement is not very effective to control the epidemic.

## 2 The model

We want to predict the daily number of observed contaminated people that we will call the positive cases because they are positive to the PCR or antigenic test. Because the testing process is not uniform in a week, we will use the weekly average of the daily positive cases as observed data. Let us denote by *y*(*t*) the cumulated number of positive cases from the epidemic start. Then we define *z*(*t*) = log(*y*(*t*) *− P*) where *P* is a number to be estimated as the cumulated number of positive cases infected by the previous variants of the one currently studied. Then we use the model *ż* = *a − bz*^*r*^ discussed in [3] where the three parameters *a, b, r* must be estimated. Before discussing a way to estimate the unknown parameters, let us give the results obtained for the French case.

In the French case see Figure-1 we can distinguish four modes : April, November 2020, May and September 2021.

**Figure 1:**
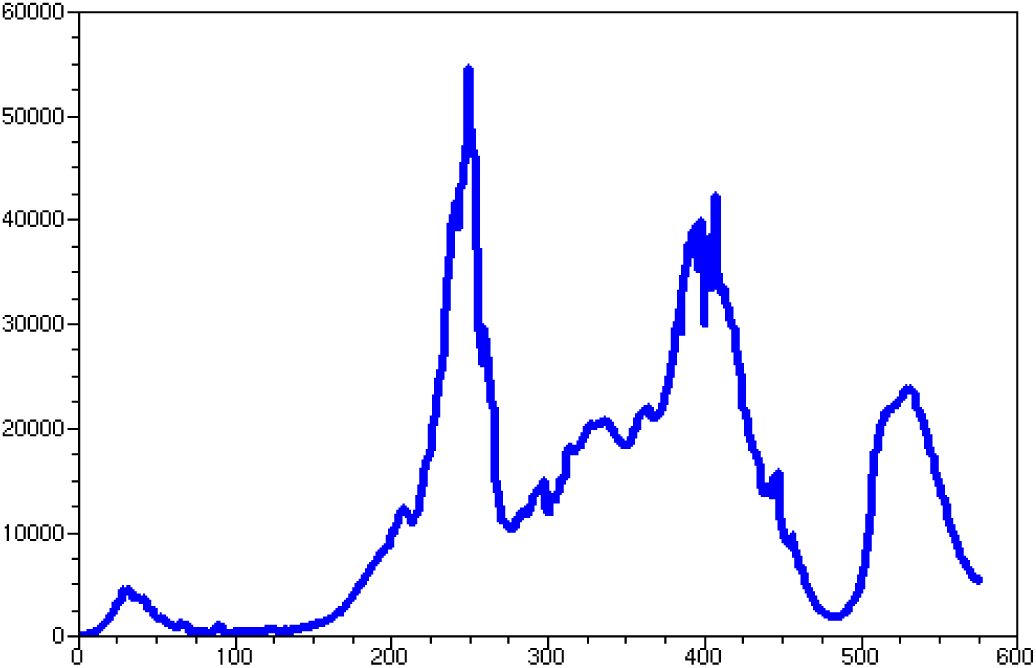
Number of the daily positive people in France (averaged on a week).

For these four modes, let us give the results obtained. We compare the average daily positive number observed with the predicted one given by the models. The complete set of data has been cut in four subsets. On each subset we have estimated the four parameters *a, b, r, P* based on data in blue or in blue and green and we have given the corresponding prediction obtained respectively in red and black Figure-2-3.

**Figure 2:**
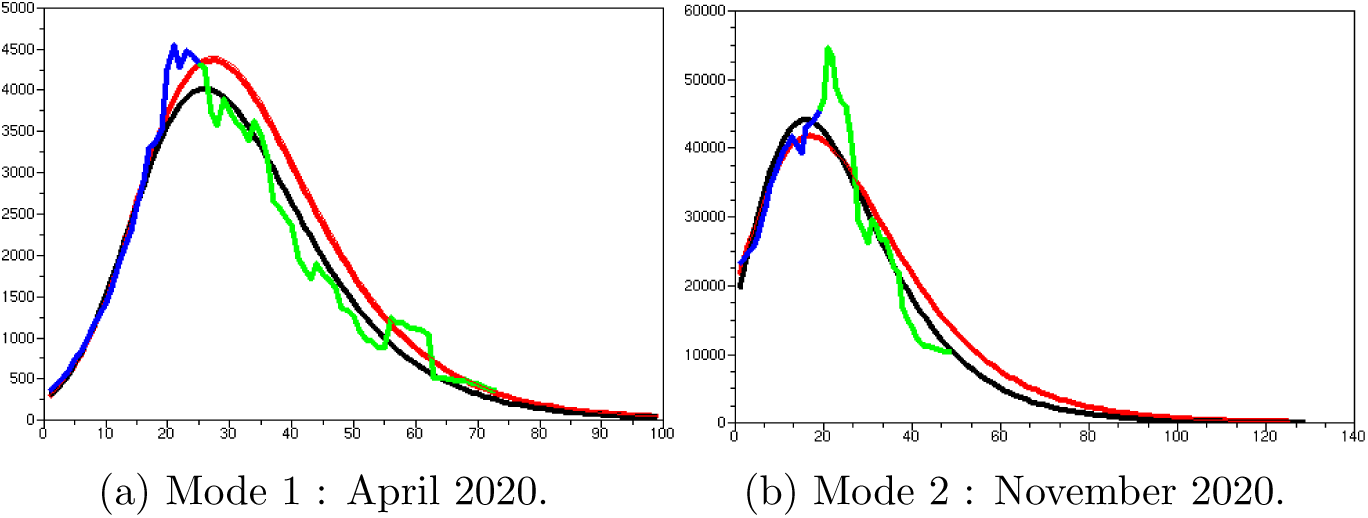
Blue and Green : observations, Red : model prediction based on blue data, Black: model prediction based on the blue and green data.

**Figure 3:**
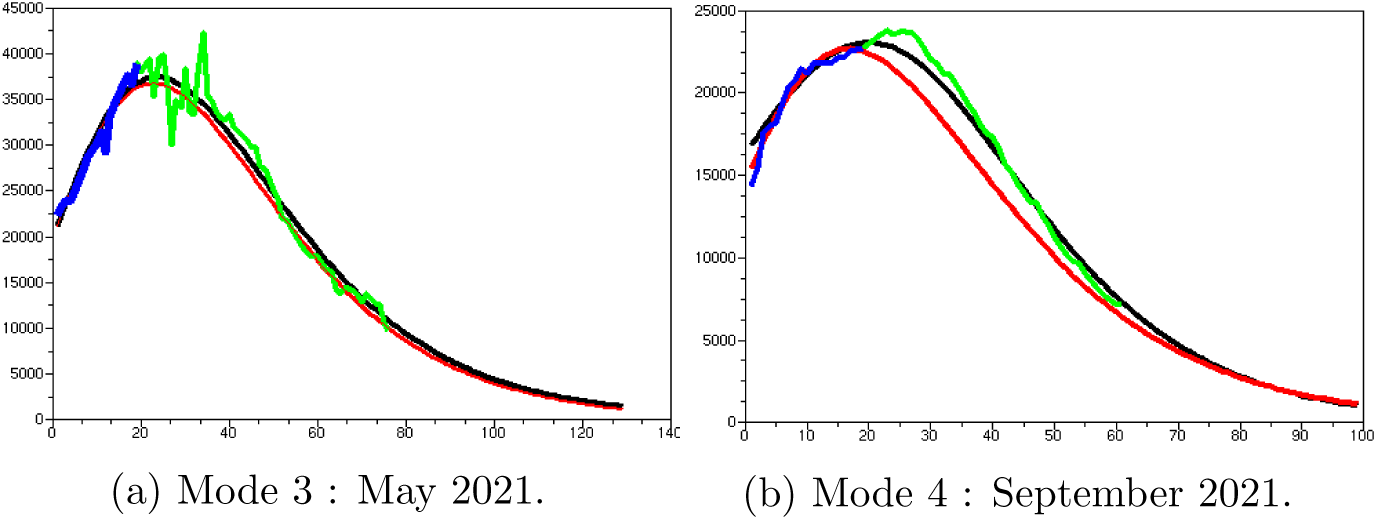
Blue and Green : observations, Red : model prediction based on blue data, Black : model prediction based on the blue and green data.

The main point of the paper.

Denoting *y*(*t*) the cumulated observed number of people infected during the Covid epidemic until time *t*, the time function :

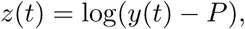

where *P* is parameter to be estimated, can be well approximated, during a mode, by a first-order time invariant stable nonlinear dynamical system:

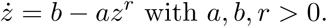

We observe that with few data (about twenty observations) before reaching its maximum, we are able to predict the epidemic decrease. But the quality of the result crucially depends on the choice of these data. A way to choose this set is to start after the inflection point in the increase speed epidemic (when we see on the daily positive cases observed data that the speed of increase begin to slow down). But we are not able with data of the epidemic start to predict the maximum of the mode. Nevertheless, for the second and third mode, the dates of the last data, in blue, used to generate the red predictions happen before the confinement decisions.

Moreover, the small difference between the red prediction based on the blue data with respect to the black prediction based on the union of the blue and green data suggests a weak impact of the confinement decisions.

## 3 Parameter Estimation

We have to estimate the four parameters *a, b, r, P*.

Given *r* and *P* it is easy to estimate the two parameters *a, b* since we observe *z*(*t*) from which we compute *ż* and a simple linear regression gives *a* and *b* from the differential equation *ż* = *b − az*^*r*^.

In [3] we have seen the qualitative role of *r* which can increase the negative slope of the daily positive cases number in the final part of the epidemic. We adapt it only when we are not able to achieve a good result with the simplest case *r* = 1.

The parameter *P* is very important and it must be optimized to obtain the best fit possible with the daily positive cases curve. For that we compute the error between the observed curve and the prediction. If it is positive we increase *P* ; if it is negative we decrease *P*. This defines a kind of median curve. The computation of the gradient is not simple and is not necessary since we have to optimize only a scalar *P*. The initial value of *P* is taken a bit smaller than the first value of *y* of the time interval considered in the mode that we are trying to predict.

Let us take the example of the third mode of the French case. The time interval considered is [375, 490] see Figure-1. Then *y*(375) = 3 882 430, we start with *P* = 3 882 000, *r* = 1 and the best linear estimators of *a* and *b*. We obtain the daily cases prediction given in the Figure-4. After the optimization of the *P* parameter (*P* = 3 330 436) we obtain the prediction given in the Figure-5.

**Figure 4:**
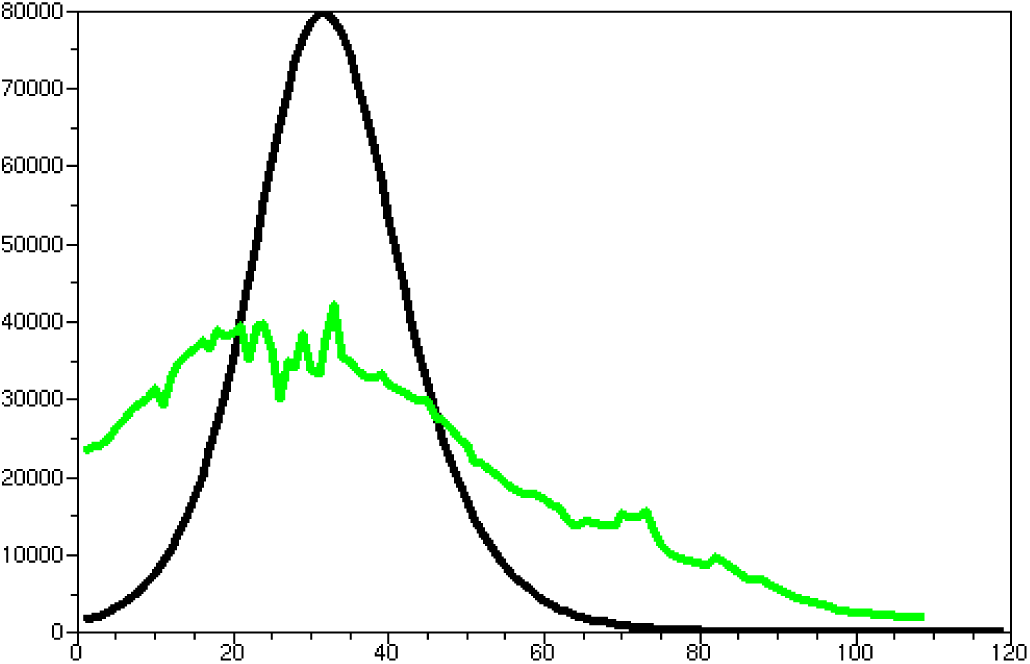
Third mode daily prediction with the initial *P*.

**Figure 5:**
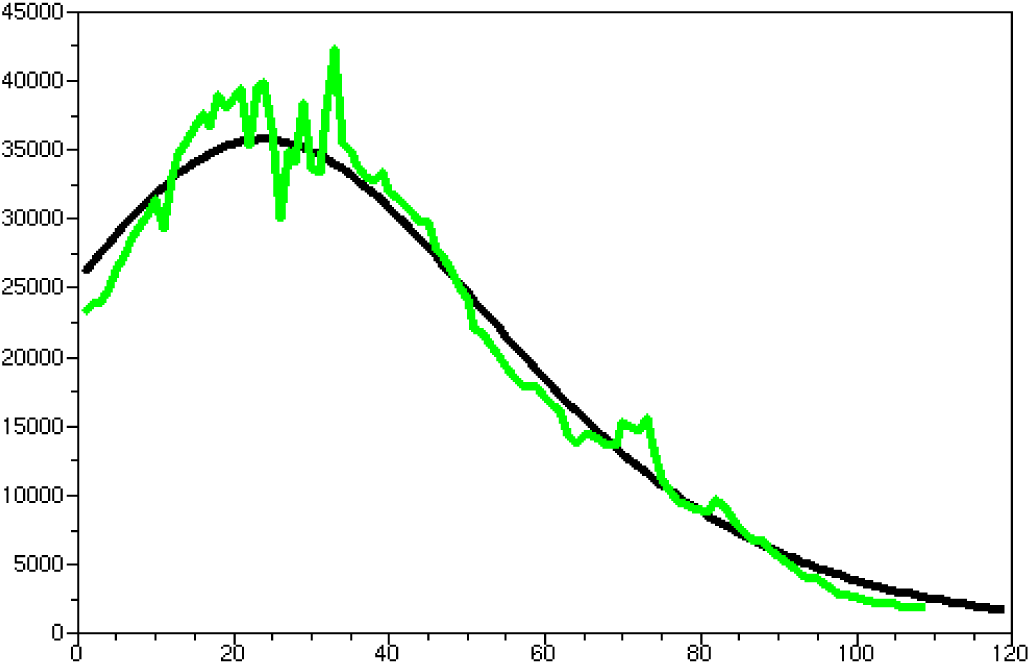
Third mode daily prediction with the optimized *P*.

We have not to tried an another *r* since the result is good. We remark the important role plaid by the parameter *P*.

## 4 Conclusion

We have shown that a correct prediction of a mode of the French Covid epidemic is provided by fitting a nonlinear time-invariant first-order dynamical model *ż* = *b − az*^*r*^ on the logarithm of the number of observed contaminated people minus a parameter *P* to be estimated. The results obtained on the four modes in the French case are satisfactory. Based on this way of predicting the epidemic modes, we also see that the confinement decisions were not very effective.

## Data Availability

All data are available at https://geodes.santepubliquefrance.fr/

